# Epidemiology of Myalgic Encephalomyelitis among individuals with self-reported Chronic Fatigue Syndrome in British Columbia, Canada, and their health-related quality of life

**DOI:** 10.1101/2024.05.16.24307437

**Authors:** Enkhzaya Chuluunbaatar-Lussier, Melody Tsai, Travis Boulter, Carola Muñoz, Kathleen Kerr, Luis Nacul

## Abstract

**Background:** There is no accurate data on the epidemiology of Myalgic Encephalomyelitis/ Chronic Fatigue Syndrome (ME/CFS) in Canada. The aims of the study were to describe the epidemiology of confirmed ME/CFS cases and their health-related quality of life (HRQoL).

**Methods:** This is a cross-sectional study with British Columbia Generations Project (BCGP) participants who self-reported having CFS and population-based controls with no fatiguing illness. Participants completed the Symptoms Assessment Questionnaire, RAND 36-item Health Survey, and Phenotyping Questionnaire Short-form. These assessments enabled the identification and characterization of “confirmed cases” of ME/CFS. Those with self-reported diagnoses who did not meet study diagnosis of ME/CFS were subcategorized as “non-ME/CFS cases.”

**Results:** We included 187 participants, 45.5% (n=85) self-reported cases and 54.5% (n=102) controls; 34% (n=29) of those who self-reported ME/CFS fulfilled diagnostic criteria for ME/CFS. The population prevalence rates were 1.1% and 0.4% for self-reported and confirmed ME/CFS cases respectively. Participants displayed significantly lower scores in all eight SF-36 domains compared to the other groups. Mental component scores were similar between ME/CFS and non-ME/CFS groups. The main risk factor for low HRQoL scores was fatigue severity (β = - 0.6, p<0.001 for physical health; β = -0.7, p<0.001 for mental health).

**Conclusions:** The majority of self-reported cases do not meet diagnostic criteria for ME/CFS, suggesting that self-reported CFS may not be a reliable indicator for a true ME/CFS diagnosis. HRQoL indicators were consistently lower in ME/CFS and non-ME/CFS cases compared to controls, with ME/CFS cases having lower scores in most domains. Having higher symptom severity scores and perceived poorer health were the significant affecting factors of lower

HRQoL. Although self-report can be used as screening to identify cases in populations, we suggest studies of ME/CFS should include appropriate medically confirmed clinical diagnosis for validity. Further large-scale population-based studies with simultaneous medical assessment are suggested to further characterize validity parameters of self-reported diagnosis.

## INTRODUCTION

Myalgic Encephalomyelitis or Chronic Fatigue Syndrome (ME/CFS) is a chronic multi-system debilitating disease characterized by intolerance to physical, cognitive, or emotional exertion. The prevalence of clinically diagnosed ME/CFS is estimated to be 0.2 to 1.4% [1, 2], while a meta-analysis (0.7%) and a pooled prevalence (0.4%) reported in the systematic review [2]. The reported estimated number of people having ME/CFS in Canada increased 37% from 407,789 in 2014 [3] to 561,500 in 2017 [4]. This apparent increase in prevalence may not be reliable, as it was based on self-reports (of being diagnosed with CFS). ME/CFS has been shown to significantly affect the quality of lives of individuals [5]. Health-related quality of life (HRQoL) is significantly lower in individuals with ME/CFS compared to the healthy population [5-7] and to those diagnosed with other long-term chronic illnesses such as multiple sclerosis [5, 8, 9] and cancer [8]. Both physical and mental HRQoL dimensions are affected in ME/CFS, though physical health is more significantly affected.

The underlying etiology of ME/CFS is not fully understood, although several etiological factors have been investigated, including infections [10, 11], neurological [12], immunological [13, 14], endocrine [15], and genetic [16]. Individuals with ME/CFS experience debilitating symptoms, with around 25% estimated to be housebound and 16% bedridden [17]. Mirin et al. estimated the economic impact to be $149 to $362 billion due to increased prevalence of ME/CFS (4-9 million individuals) in the United States after the COVID-19 pandemic [18]. There are no definitive diagnostic tests nor evidence of effective drug treatments to provide patients affected by ME/CFS with a cure. However, new treatments are emerging [19-21], and with proper and early diagnosis and management, there is an increased likelihood of improvement or reversal of symptoms [20, 22].

Currently, there is no accurate data on the health status of patients with ME/CFS from across Canada. In this study, we aimed to describe the epidemiology of ME/CFS among adults in the province of British Columbia (BC), investigate the HRQoL and associated risk factors, and assess feasibility of confirming ME/CFS cases among self-reported population. Our main hypotheses are that the population prevalence of ME/CFS in BC is similar or larger than that estimated internationally, i.e. we expect an estimated point prevalence rate to be around 0.4% (often quoted as an international average rate) [2]. We also expect age variation and higher prevalence in women. The study served to assess the feasibility of using the methodology in larger populations in Canada.

## METHODS

This population-based cross-sectional study, nested within a prospective cohort study, was undertaken as part of a Canadian Institutes of Health Research (CIHR) funded network application for the Interdisciplinary Canadian Collaborative Myalgic Encephalomyelitis (ICanCME) Network, a national interdisciplinary collaborative ME/CFS research network that focuses on patient-centered research, from discovery to implementation. The goal of the network is to advance research in finding cause(s) and possible treatments for ME/CFS, thereby reducing the impact of ME/CFS on the health of Canadians. The understanding of the epidemiology of ME/CFS is essential to inform service delivery for this population.

### Study Population and Cohort

Participants were recruited from the BC Generations Project (BCGP) [23], the British Columbian regional cohort of the Canadian Partnership for Tomorrow’s Health (CanPath) project [24]. CanPath is a population-based cohort looking at the social, economic and health data of over 330,000 Canadians, aged between 30 and 74 years at recruitment. The cohort includes participants from Alberta (n∼55,000), BC (n∼30,000), Manitoba (n∼10,000), Quebec (n∼43,000), Ontario (n∼225,000) and the Atlantic provinces (n∼36,000). The BCGP recruited new participants between October 2009 and March 2013 (“1^st^ participant time-point” at baseline), with most participants having completed physical measurements, Health & Lifestyle Questionnaires (HLQ), and had biological samples (blood and urine) (Table 1). In September 2016, during the “2^nd^ participant time-point”, a follow-up HLQ was released to all participants to collect any updates, including a newly added section asking if they were ever diagnosed with Chronic Fatigue Syndrome (CFS). We do not know the reliability of a self-reported diagnosis of CFS, and particularly, what proportion of them meet the rigorous diagnostic criteria for ME/CFS. Therefore, we considered those with a “self-reported” diagnosis as “*probable cases.*” Control individuals were randomly selected from the BCGP database and contributed to estimates of “willingness of individuals without ME/CFS or other fatiguing illnesses to participate in ME-specific future studies” and investigation of risk factors.

**Table 1.** Data collection timeframe.

|  | 1<sup>st</sup> Time-point<br>(BCGP) | 2<sup>nd</sup> Time-point<br>(BCGP) | 3<sup>rd</sup> Time-point<br>(BC EP-ME) |
| --- | --- | --- | --- |
| Health & Lifestyle Questionnaires (HLQ) | X | X* |  |
| Physical measurements | X |  |  |
| Symptoms Assessment Questionnaire (SAQ) |  |  | X |
| RAND 36-Item Short Form Health Survey (SF-36) |  |  | X |
| Phenotyping Questionnaire Short Form (pQsym-12) |  |  | X |

Note: \* Including the self-report having chronic fatigue syndrome

### Data Collection and Questionnaires

In 2016, at the 2^nd^ participant time-point, 19,145 participants responded to the question about their Chronic Fatigue Syndrome diagnosis. Of those participants, 217 (1.1%) responded ‘yes’ to ever having been diagnosed with CFS. Participants were given the option to consent to being contacted for future studies. Thus, almost 90% (195 out of 217) of BCGP participants who self-reported CFS and had previously consented to being contacted for future studies were re-contacted and invited to participate in this study. All participants were invited to complete the same questionnaires which corresponded to the “3^rd^ participant time-point” (Table 1). More details surrounding recruitment are shown in S1 Figure.

Consenting individuals completed the following three questionnaires: 1) Symptoms Assessment Questionnaire (SAQ) for diagnosis confirmation, which is a symptom specific questionnaire for ME/CFS diagnosis aid [25]; 2) Phenotyping Questionnaire Short Form (PQsym-12), a symptom severity assessment with 12-questions created by the CureME Group in the United Kingdom used to collect data enabling clinical phenotyping information, with higher scores representing greater severity of ME/CFS symptoms [25]; and 3) RAND 36-Item Short Form Health Survey (SF-36) to assess physical and mental function, with scores ranging between 0-100 for eight domains, and two summary normalized scores for overall physical and mental functions, with higher scores representing better health [26].

### Case Definitions of ME/CFS

In the absence of biomarkers for diagnosis, case definitions of ME/CFS were based on symptoms presented by participants. Responses to the SAQ enabled the identification of “confirmed cases” of ME (categorized as the “ME/CFS group”), as it contains a built-in algorithm that enables categorization of participants according to whether they meet the diagnostic criteria of the 2003 Canadian Consensus Criteria (2003 CCC) [27] and/or 2015 Institute of Medicine (2015 IOM) criteria [28]. Those with a self-reported diagnosis, but who do not meet the study diagnosis of ME, were sub-categorized as the “non-ME/CFS group.”

### Data Analysis

All analyses were performed using Stata17 statistical packages [29]. Descriptive analyses for socio-demographic variables, sleep, and symptom severity scores (PQSym-12) are presented as percentages with 95% confidence interval (CI) for categorical variables, and means and 95% CI for continuous variables. Normal distributions were verified using histograms, box plots or scatter plots for continuous variables such as age, sleep hours, symptom severity scores, and all eight domains of SF-36 scores. Mean scores of each health domain of SF-36, as well as summary physical and mental component scores, were calculated based on the user manual created by Ware et al.[26, 30]. Scoring for each domain included precoding numeric values for each question, then averaging to eight domains with resulting scores ranging from 0-100 [30]. To calculate physical and mental component summary scores, we first performed z-score transformation to standardize each domain score by using the standard deviation from the US general population, followed by aggregating scale scores for physical and mental components, and lastly, calculating physical and mental summary scores accordingly [31]. One-way ANOVA was performed for comparing physical and mental summary components of SF-36 across groups. The post-hoc Tukey methods (ME/CFS vs Non-ME/CFS; ME/CFS vs. Control; and Non-ME/CFS vs. Control) was employed for all eight domain scores, and physical and mental component scores. Unadjusted and adjusted linear regression analyses were performed to evaluate the relationship between factors associated with HRQoL, with physical and mental summary component scores as dependent variables. Two independent models, one for physical and one for mental summary scores, were created. Diagnosis group (ME/CFS, non-ME/CFS and controls) was treated as a main exposure for both models. Independent variables included age, sex, education level, ethnicity, employment status, sleep hours, sleeping trouble, and PQSym-12 scores. Models were adjusted for selected factors significantly associated with both outcome and main exposure in unadjusted model. Moreover, for some of the analyses, ME/CFS and non-ME/CFS groups were combined as “*probable cases*” and compared to the controls; this was due to the relatively small sample sizes for both of the groups. Lastly, comparison between ME/CFS and non-ME/CFS was performed for both physical and mental health summary scores.

## RESULTS

### Prevalence rates and descriptive epidemiology

The prevalence of self-reported CFS was 1.1% (217 out of 19,145) at ascertained at the 2^nd^ participant time-point. To determine confirmed ME/CFS prevalence, a total of 700 BCGP participants (195 probable cases and 505 healthy controls) were invited to participate in this study. Response rates, defined as the percentage of participants who completed all three questionnaires, were 43.6% (n=85) among probable cases and 20.2% (n=102) in the control group, respectively, with a total of 187 participants. Among the 85 individuals who reported having CFS, 34% (n=29) fulfilled the IOM and/or CCC diagnostic criteria, yielding a prevalence for confirmed ME/CFS diagnosis of 0.4%. These individuals were categorized into the ME/CFS group, while the remaining 56 (66%) individuals were categorized into the non-ME/CFS group (Table 2).

**Table 2.**
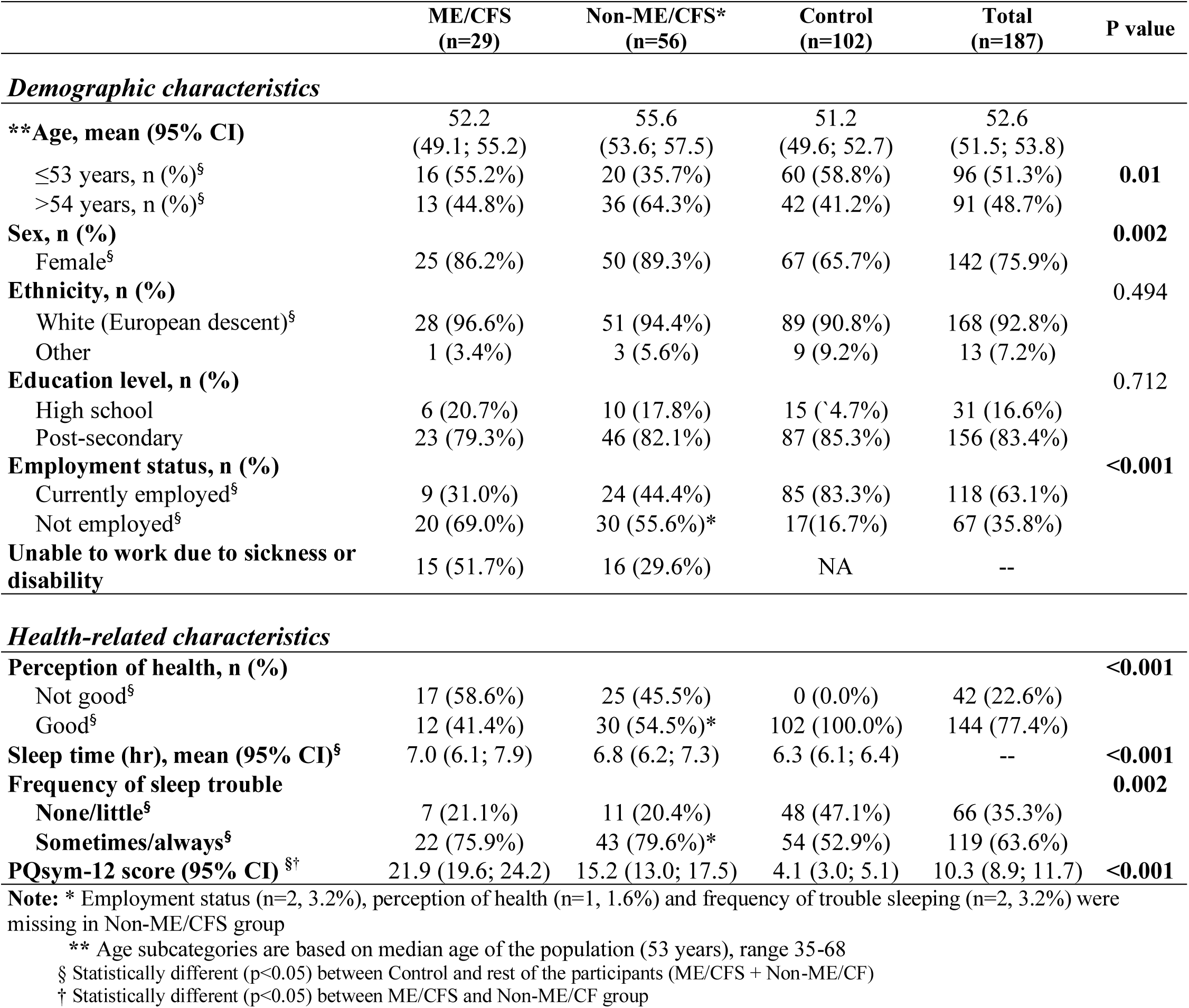
Descriptive characteristics of the study participants.

Age of the study population ranged between 35 and 68 years, with a mean age of the total population of 52.6 years (95% CI 51.5; 53.8). Those in the non-ME/CFS group (55.6 years, 95% CI 53.6; 57.5) were slightly older than ME/CFS participants (52.2 years, 95% CI 49.1; 55.2), and were significantly older than the control participants (51.2 years, 95% CI 49.6; 52.7, p = 0.01). Participants were mostly female (86.2% in ME/CFS, 89.3% in non-ME/CFS, and 65.7% in control groups). More than 92% of the participants identified as “white” ethnicity. Education level in the three groups was not significantly different; however, more than 80% of the study population had post-secondary education. Almost 70% (n=20) of the ME/CFS group and 56% (n=30) of non-ME/CFS were unemployed. Among them, 56% (n=15) and 30% (n=16) reported physical health conditions as the reasons for unemployment in the ME/CFS and non-ME/CFS groups, respectively. Only 17% (n=17) of controls were unemployed.

More than half (58.6%) of the individuals from the ME/CFS group and just under half (45.5%) of non-ME/CFS group reported poor or fair health (categorized as ‘not good’). In contrast, none of control participants reported poor or fair health (p < 0.001). Moreover, although individuals in the ME/CFS group reported sleeping significantly longer hours than other groups, 76% of ME/CFS and 80% of non-ME/CFS groups had trouble sleeping, compared to 53% in the control group (p=0.002). The outliers (n=3) in sleep hours were removed from further sleep hours related analysis, and differences of these continuous variables were tested in three groups using one-way ANOVA (ME/CFS, non-ME/CFS and controls).

The PQSym-12 questionnaire results showed that the ME/CFS group had significantly higher symptom severity scores (21.9, 95% CI 19.6; 24.2), when compared to both the non-ME/CFS (15.2, 95%CI 13.0; 17.5, p<0.001) and control (4.1, 95% CI 3.0; 5.1, p<0.001) groups (Table 2). When comparing the three groups, each group was statistically significant different from the other two groups (see also Figure 1).

**Figure 1.**
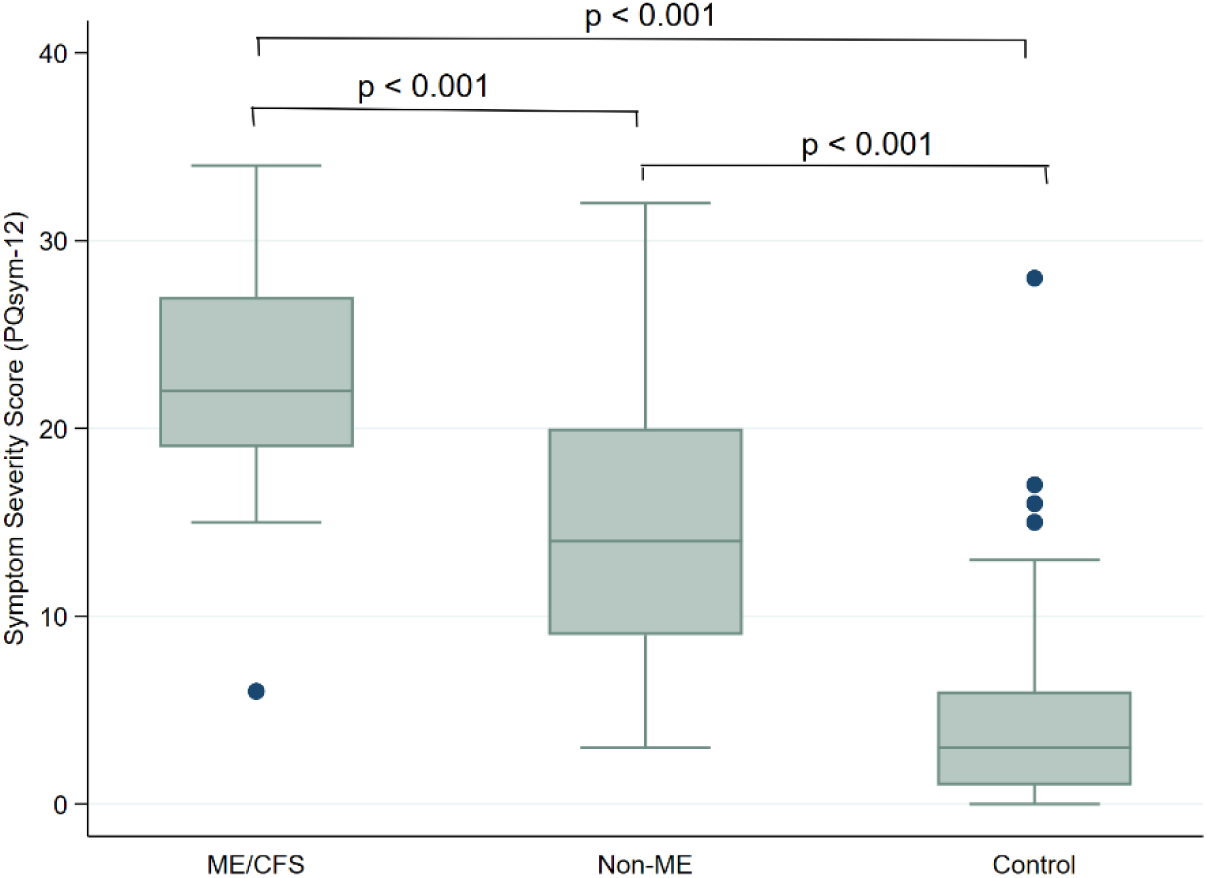
Symptom Severity Scores in three groups.

### Health-Related Quality of Life

HRQoL SF-36 scores are shown in Table 3. Domain scores were presented as average scores from corresponding questions, while physical and mental component summary scores were transformed as discussed in the Methods section. For all eight domain scores, the scores were significantly different across the three groups, with the ME/CFS group consistently showing the lowest values (except for emotional well-being) and the control group without fatigue had the highest. ANOVA post-hoc Tukey results showed that compared to non-ME/CFS participants, ME/CFS participants had lower scores in physical functioning (p<0.001), role limitations due to physical health (p=0.023), pain (p=0.008), energy/fatigue (p=0.018), and social functioning (p=0.001) domains and physical health component score (p<0.001). There were no significant differences in role limitation due to emotional problems, emotional wellbeing, and general health domains as well as mental health component scores between ME/CFS and non-ME/CFS groups. In contrast, both groups had significantly lower HRQoL scores than controls in all eight domains and the two summary scores (p<0.001). This was more pronounced in the role limitation due to physical functioning domain (5.9 in ME/CFS vs. 23.2 in non-ME/CFS, and 87.0 in controls, p<0.001). When comparing physical summary scores between the three groups (Fig 2a), there was a significant difference between ME/CFS and non-ME/CFS groups (p=0.001), and a highly significant difference between ME/CFS and controls (p<0.001), and non-ME/CFS and controls (p<0.001) group. Mental component scores were similar in ME/CFS and non-ME/CFS groups (p=0.613), but both ME/CFS and non-ME/CFS groups had significantly lower scores than the control group (both p<0.001) (Fig 2b).

**Figure 2a.**
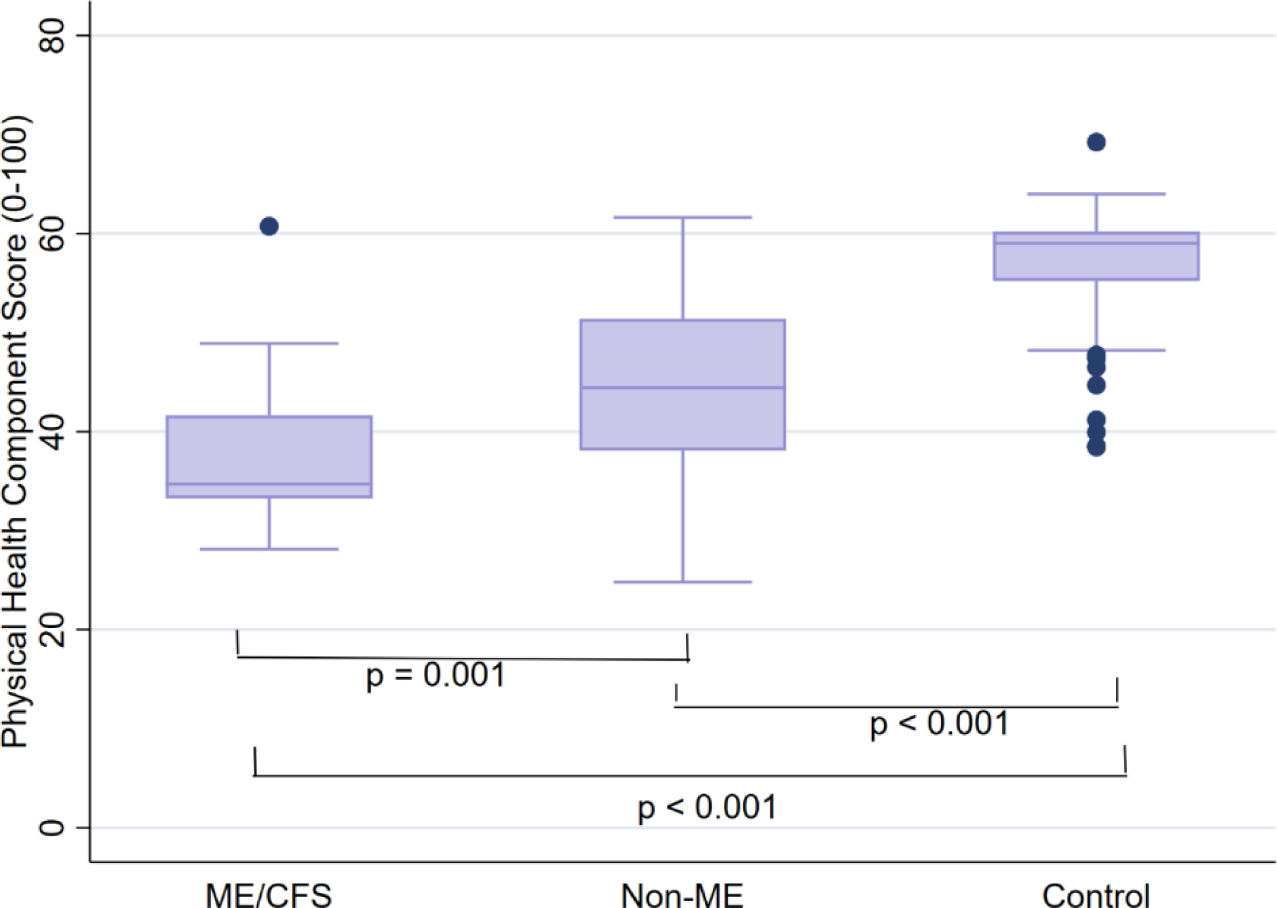
Physical Health Component Scores (SF-36) in three Groups.

**Figure 2b.**
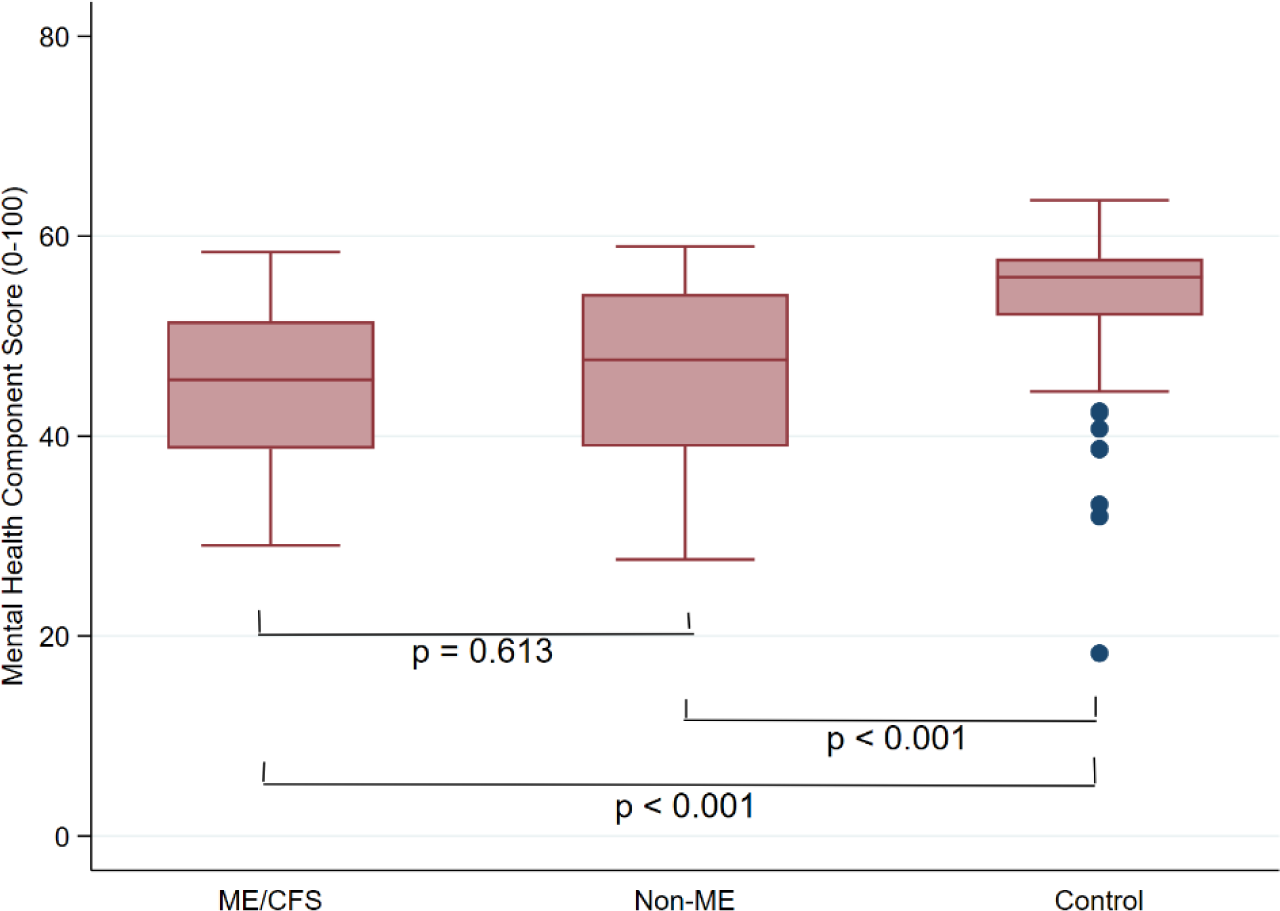
Mental Health Component Scores (SF-36) in three groups.

**Table 3.**
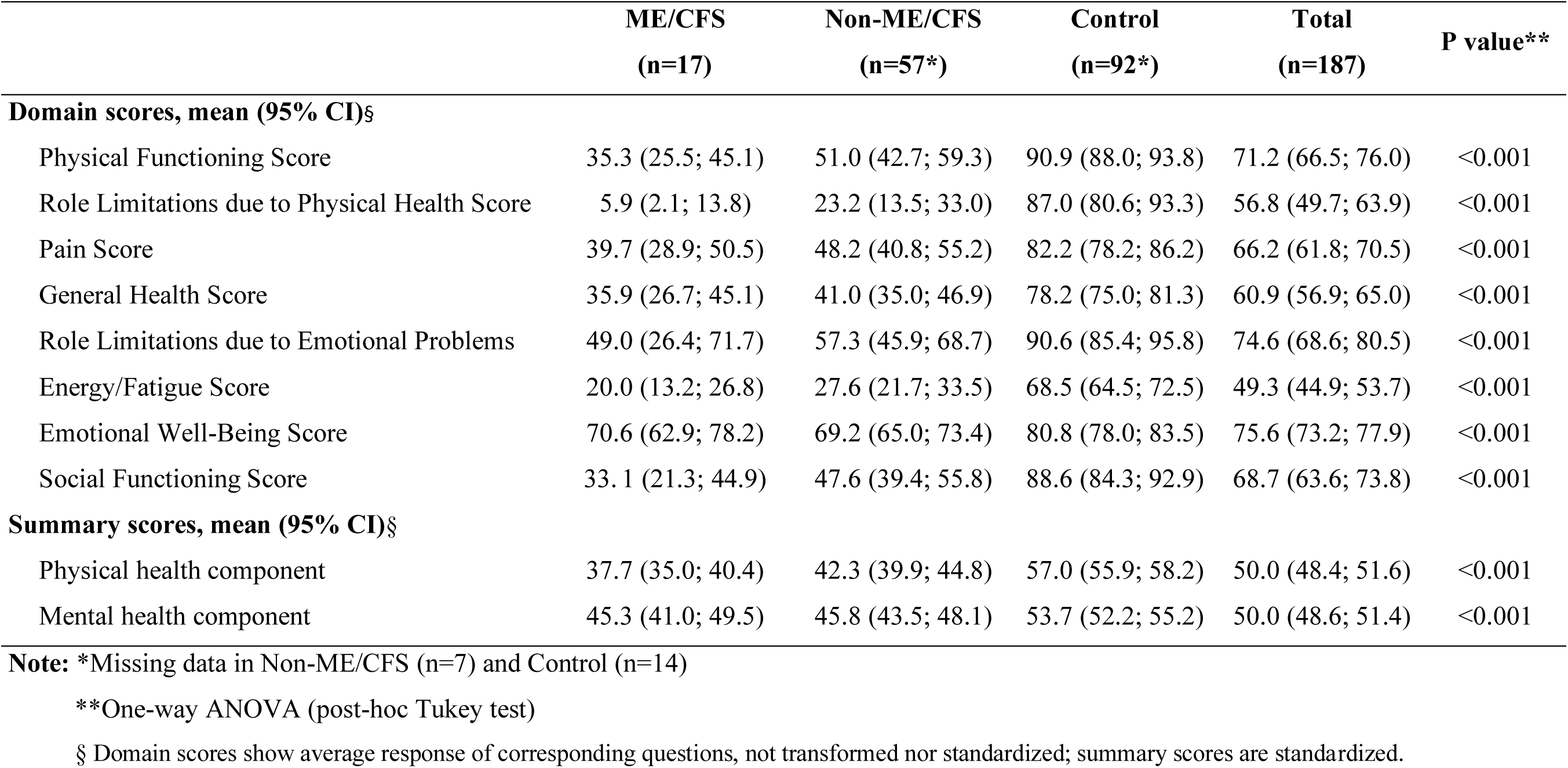
SF-36 domain scores (higher score implicates less impairment)

|  | ME/CFS<br>(n=17) | Non-ME/CFS<br>(n=57*) | Control<br>(n=92*) | Total<br>(n=187) | P value** |
| --- | --- | --- | --- | --- | --- |
| **Domain scores, mean (95% CI)§** |  |  |  |  |  |
| Physical Functioning Score | 35.3 (25.5; 45.1) | 51.0 (42.7; 59.3) | 90.9 (88.0; 93.8) | 71.2 (66.5; 76.0) | <0.001 |
| Role Limitations due to Physical Health Score | 5.9 (2.1; 13.8) | 23.2 (13.5; 33.0) | 87.0 (80.6; 93.3) | 56.8 (49.7; 63.9) | <0.001 |
| Pain Score | 39.7 (28.9; 50.5) | 48.2 (40.8; 55.2) | 82.2 (78.2; 86.2) | 66.2 (61.8; 70.5) | <0.001 |
| General Health Score | 35.9 (26.7; 45.1) | 41.0 (35.0; 46.9) | 78.2 (75.0; 81.3) | 60.9 (56.9; 65.0) | <0.001 |
| Role Limitations due to Emotional Problems | 49.0 (26.4; 71.7) | 57.3 (45.9; 68.7) | 90.6 (85.4; 95.8) | 74.6 (68.6; 80.5) | <0.001 |
| Energy/Fatigue Score | 20.0 (13.2; 26.8) | 27.6 (21.7; 33.5) | 68.5 (64.5; 72.5) | 49.3 (44.9; 53.7) | <0.001 |
| Emotional Well-Being Score | 70.6 (62.9; 78.2) | 69.2 (65.0; 73.4) | 80.8 (78.0; 83.5) | 75.6 (73.2; 77.9) | <0.001 |
| Social Functioning Score | 33.1 (21.3; 44.9) | 47.6 (39.4; 55.8) | 88.6 (84.3; 92.9) | 68.7 (63.6; 73.8) | <0.001 |
| **Summary scores, mean (95% CI)§** |  |  |  |  |  |
| Physical health component | 37.7 (35.0; 40.4) | 42.3 (39.9; 44.8) | 57.0 (55.9; 58.2) | 50.0 (48.4; 51.6) | <0.001 |
| Mental health component | 45.3 (41.0; 49.5) | 45.8 (43.5; 48.1) | 53.7 (52.2; 55.2) | 50.0 (48.6; 51.4) | <0.001 |

**Note:** \*Missing data in Non-ME/CFS (n=7) and Control (n=14)
\*\*One-way ANOVA (post-hoc Tukey test)
§ Domain scores show average response of corresponding questions, not transformed nor standardized; summary scores are standardized.

#### Risk factors for Poor Physical Health Summary Score

Univariate analyses (unadjusted estimates) show that compared to controls, both ME/CFS (β = -19.9, p<0.001) and non-ME/CFS (β = -13.0, p<0.001) groups have significantly lower physical health summary scores (Table 4). Other factors inversely associated with physical health were female sex (β = -5.3, p=0.003), not in employment (β = -10.0, p<0.001), perception of general health (β = -17.4, p<0.001), frequency of sleep trouble (β = -6.0, p<0.001), and symptom severity (β = -1.0, p<0.001). The correlation analysis of main outcomes showed strong correlations between PCS and both symptom severity score (by PQSym-12) and general health. (Table 5). The adjusted model showed that compared to the control group, both ME/CFS and non-ME/CFS had significantly low physical health scores when adjusting for age, sex, education, ethnicity, and employment. Compared to employed people, who had been not-employed had higher physical health scores (β = 3.2 p=0.02) after adjusting for other covariates (Table 6a).

**Table 4.** Univariate regression analysis for Physical and Mental component scores.

| Groups | Physical component (n = 166) |  |  | Mental component (n = 166) |  |  |
| --- | --- | --- | --- | --- | --- | --- |
|  | Estimates | 95% CI | P value | Estimates | 95% CI | P value |
| **Control - reference** | 57.1 | 55.9; 58.3 |  | 53.7 | 52.0; 55.3 |  |
| Non-ME/CFS | -13.0 | -15.9; -10.1 | **<0.001** | -7.4 | -10.5; -4.4 | **<0.001** |
| ME/CFS | -19.9 | -22.6; -17.2 | **<0.001** | -8.5 | -11.9; -5.1 | **<0.001** |
| **Age** | -0.1 | -0.3; 0.1 | 0.263 | 0.1 | -0.04; 0.3 | 0.129 |
| **Sex (ref: male)** | 54.1 | 51.2; 57.0 |  |  |  |  |
| Female | -5.3 | -8.8; -1.8 | **0.003** | -2.4 | -5.9; 1.1 | 0.170 |
| **Ethnicity (ref: other than White)** | 55.2 | 47.7; 62.7 |  |  |  |  |
| White | -5.3 | -13.0; 2.3 | 0.172 | -4.5 | -8.5; -0.5 | **0.028** |
| **Education (ref: high school)** | 46.7 | 42.4; 50.9 |  |  |  |  |
| Post-secondary | 4.1 | -0.6; 8.7 | 0.085 | 3.4 | -0.1; 7.1 | 0.061 |
| **Employment (ref: employed)** | 53.7 | 52.0; 55.5 |  |  |  |  |
| Not employed | -10.0 | -13.1; -6.8 | **<0.001** | -1.5 | -4.4; 1.3 | 0.291 |
| **Perception of health (ref: good)** | 54.1 | 52.7; 55.5 |  |  |  |  |
| Not good | -17.4 | -20.0; -14.9 | **<0.001** | -7.4 | -10.5; -4.2 | **<0.001** |
| **Sleep hours/day** | -1.1 | -2.2; -0.1 | 0.064 | -0.55 | -1.5; 0.4 | 0.263 |
| **Sleep trouble (ref: none/ little)** | 53.8 | 51.5; 56.1 |  |  |  |  |
| Sometimes/always | -6.0 | -9.1; -2.8 | **<0.001** | -2.3 | -5.1; 0.4 | 0.098 |
| **PQSym-12 score** | -1.0 | -1.1; -0.9 | **<0.001** | -0.6 | -0.7; -0.5 | **<0.001** |

**Note:** Ethnicity and education were excluded from the adjusted model due to large number of missing values or outliers. PQSym-12 score – symptom severity score

**Table 5.**
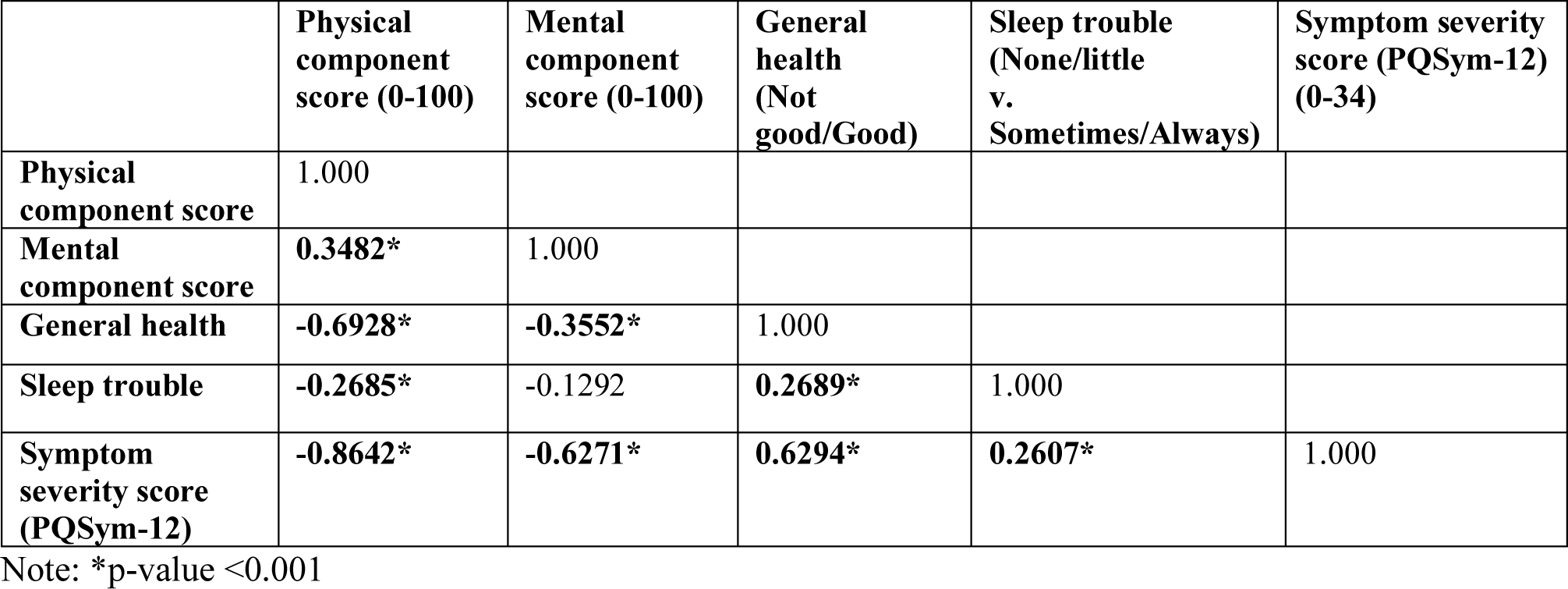
Correlations among key study outcomes, n=161.

**Table 6a.** Adjusted regression model for Physical Health.

| Variables | Adjusted Model (n=160) |  |  |
| --- | --- | --- | --- |
|  | Estimates | 95% CI | P value |
| **Groups** |  |  |  |
| Control (ref) | 52.9 | 44.7; 61.0 | **<0.001** |
| Non-ME/CFS | -11.7 | -15.2; -8.2 | **<0.001** |
| ME/CFS | -18.3 | -21.2; -15.3 | **<0.001** |
| Age | 0.1 | -0.04; 0.2 | 0.186 |
| Sex (ref-male) |  |  |  |
| Female | -0.3 | -2.6; 2.0 | 0.783 |
| Ethnicity (ref-other than white) |  |  |  |
| White | -1.2 | -4.3; 1.8 | 0.430 |
| Education (ref-high school) |  |  |  |
| Above high school | 2.2 | -1.0; 5.3 | 0.175 |
| Employment (ref – employed) |  |  |  |
| Not -employed | 3.1 | 0.5; 5.7 | **0.02** |

Note: Model does not include General Health, Sleep Trouble, and PQSym-12

**Table 6b.**
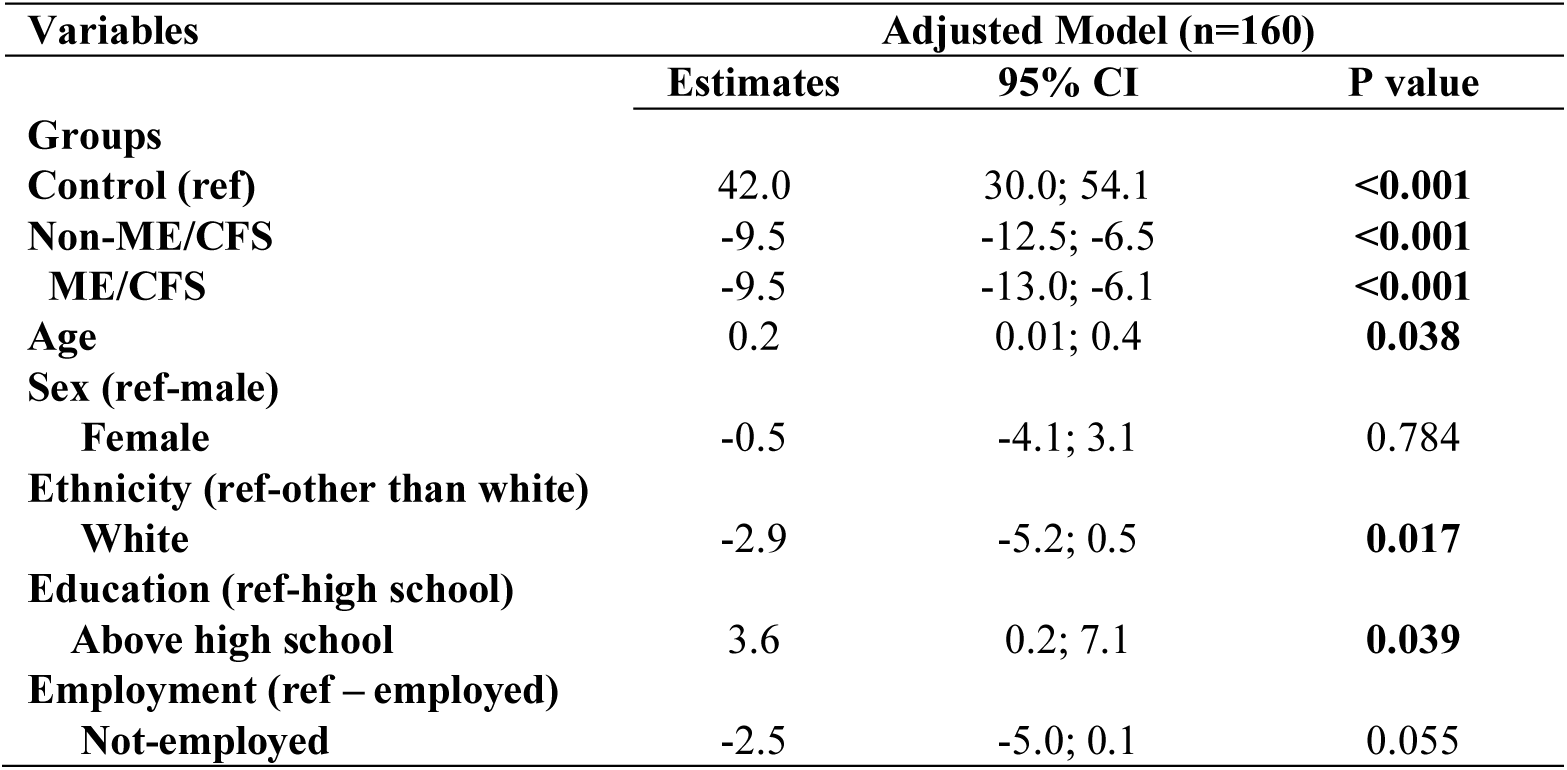
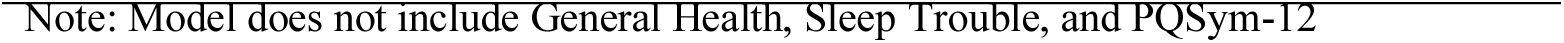
Adjusted regression model for Mental Health.

| Variables | Adjusted Model (n=160) |  |  |
| --- | --- | --- | --- |
|  | Estimates | 95% CI | P value |
| **Groups** |  |  |  |
| Control (ref) | 42.0 | 30.0; 54.1 | **<0.001** |
| Non-ME/CFS | -9.5 | -12.5; -6.5 | **<0.001** |
| ME/CFS | -9.5 | -13.0; -6.1 | **<0.001** |
| Age | 0.2 | 0.01; 0.4 | **0.038** |
| **Sex (ref-male)** |  |  |  |
| Female | -0.5 | -4.1; 3.1 | 0.784 |
| **Ethnicity (ref-other than white)** |  |  |  |
| White | -2.9 | -5.2; 0.5 | **0.017** |
| **Education (ref-high school)** |  |  |  |
| Above high school | 3.6 | 0.2; 7.1 | **0.039** |
| **Employment (ref – employed)** |  |  |  |
| Not-employed | -2.5 | -5.0; 0.1 | 0.055 |

#### Mental Health Summary Score

In unadjusted estimates, both ME/CFS (β = -8.5, p<0.001) and non-ME/CFS (β = -7.4, p<0.001) groups had significantly lower mental health scores when compared to the control group (Table 4). However, when comparing ME/CFS and non-ME/CFS groups, there were no significant differences in mental health score (Fig 2b). Reports of being European (white) ethnicity (β = -4.5, p = 0.028), poor or fair health (β = -7.4, p<0.001), and high symptom severity score (β = -0.6, p<0.001) were associated with lower mental health summary scores in both ME/CFS and non-ME/CFS cases, compared to controls (Table 4). As done for physical health analysis, the final multivariate analysis excluded highly correlated variables from the regression analysis (Table 5). Compared to the control group, both ME/CFS and non-ME/CFS had significantly lower mental health scores when adjusting for age, sex, education, ethnicity, and employment. When age increased by 1-year, mental health score increased by 0.2 (p=0.038); compared to non-White people, White people had significantly lower mental health scores (β = - 2.9, p=0.017); while higher education was associated with higher mental health scores (β = 3.6, p=0.039) when controlling for other variables (Table 6b).

## DISCUSSION

This study aimed to investigate the epidemiology of ME/CFS in an existing population-based cohort database. The prevalence of ME/CFS, ascertained based on case definitions of the IOM 2015 and/or CCC 2003, was 34% among those who self-reported CFS. Self-reported CFS represented 1.13% of the BCGP cohort, suggesting a population prevalence of ME/CFS of 0.38%, which is similar to prevalence rates suggested internationally pre-covid, albeit lower than those previously suggested for Canada [3, 4]. However, this should be regarded as minimum prevalence, for various reasons. Firstly, BCGP did not count people who might have had undiagnosed ME/CFS, and therefore did not self-report as having CFS, which likely amount to significant number of people [21, 28]. The self-report of CFS was based on a survey that preceded the assessment of patients for ME/CFS. It is possible that people who would have met the criteria for ME/CFS at the time of self-report (2^nd^ time-point) no longer met them at the time of the 3^rd^ participant time-point, or vice versa, though the numbers here are likely to be low. In addition, the distribution of the population in relation to demographic characteristics may have affected the overall estimates, noting younger populations were not represented as were those of moreadvanced age. A main point to note, however, is that self-reported CFS is not reliable method of estimating prevalence of ME/CFS.

Some factors may explain diagnosis over-estimation. Over twenty diagnostic criteria for ME/CFS have been created [32]. Almost half (48.1%) of our BCGP sample were diagnosed prior to the year 2003. We do not have information on what case definitions health professionals used for their diagnoses, if any. While the CCC 2003 and IOM 2015 are currently often used as the main diagnostic criteria, many older case definitions may have been used in other criteria including the Australia 1990, Oxford 1991, Center for Disease Control and Prevention (CDC) 1994, and London 1994 [33, 34], which may represent an alternate populations, and overall tend to over-estimte actual prevalence rates. For instance, the CDC 1994 criteria is less strict compared to the CCC 2003 and the IOM 2015, as it does not include post-exertional malaise and some other symptoms as compulsory [28]. Nacul et al. (2011) and Jason et al. (2013) both noted that over half of patients who fulfill the CDC 1994 [1] definition also fulfill the CCC 2003 criteria, Nacul et al. (58%) and Jason et al. (∼75%), respectively [1, 35]. However, virtually all of those meeting the CCC 2003 will also meet the CDC 1994 criteria. Moreover, case definition and diagnosis were not robust prior to these criteria. A survey conducted in 2002 on general practitioners’ attitudes and knowledge towards ME/CFS showed almost half of the practitioners were not confident with making a diagnosis of ME/CFS nor in its treatment [36] as well as stated in Nacul et al. (2011) [1].

In addition, a systematic review conducted by Cairns et al. [37] and as reported by Nacul et al. [38] noted that although full recovery for ME/CFS is uncommon, improvement in symptoms is more commonly reported [37, 38]. Improvement rate ranged from 8-63%, depending on treatment and length of follow-up [37, 39, 40]. In the case of ME/CFS, improvement does not mean patients are free of disease, although some of them might no longer meet the diagnostic criteria of ME/CFS. All these results combined can help to explain why only 34% of self-reported cases met diagnostic criteria in our study, since self-reported diagnoses were collected retrospectively.

As expected, the majority of ME/CFS individuals in our study were female (86%). While most cases reported in the literature occur in female [28], ME/CFS is not an exclusively women’s disease. A large-scale study of medical insurance claims data in the US showed that 35-40% of patients with a ME/CFS diagnosis were men [41]. However, data from this study was from an insurance-based (Medicare) database, and thus could be biased to include the working population, which may over-represent the overall male population. Our study population had a large proportion of participants who were unemployed (∼35%), which differs from the Valdez et al. study cohort and could partially explain the discrepancy in our findings.

We also hypothesized that prevalence of ME/CFS changes with age. Our study showed about 55% of the ME/CFS cases (16 out of 29) were younger than median age of the total study population (53 years). According to the 2019/2020 Canadian Health Survey on Seniors, 3.0% of women and 1.5% of males aged over 65 years old reported being diagnosed with ME/CFS [42]. This difference in age distribution could be explained by the small number of older age groups, 95% of our study population were aged between 51 and 54 years old at the 3^rd^ participant time-point (at questionnaire completion), with only a few people (17%, n=32) in our cohort aged over 60 years.

Our findings showed that 92.8% our participants identified as White of European descent, compared to 83.5% in the BCGP, and lower values in the general population, which may be due to under-representation of ethnic minorities, rather than higher risk in the white population. The questionnaires conducted by the BCGP were only offered in English and French, therefore limiting the diversity of the cohort. Several barriers may present at every stage in health seeking behavior and the management of ME/CFS in minority groups, including lack of awareness of ME/CFS, religious beliefs, expectations of family roles, language barriers, and stereotypical beliefs [43].

HRQoL in people in both ME/CFS and non-ME/CFS groups were significantly lower than in healthy controls in all health domains. In both groups, physical and mental health were compromised, however, the ME/CFS group had more debilitating physical health. These findings were similar to the findings from previous studies, which compared HRQoL among individuals with ME/CFS, chronic illnesses and healthy populations [5, 6, 9, 44-47]. Brown et al. noted that individuals who no longer have CFS were still more disabled and symptomatic than healthy controls, and they did not return to normal levels of functionality [47]. When comparing HRQoL between ME/CFS and non-ME/CFS, all domain scores tended to be lower in the ME/CFS group, with physical functioning shown to be significantly lower. Summary mental component scores were similar in both groups, indicating the mental health of individuals who self-reported CFS was affected as much as people with ME/CFS. This confirms the physically disabling nature of ME/CFS [5] and that the mental health impact of ME/CFS is similar to that experienced by people with chronic fatigue of other causes.

Similar to our findings, a clinical-based cross-sectional study found that, compared to people with other fatiguing symptoms, people with ME/CFS had a lower index of physical role functioning and social functioning, lower energy, worse pain and poorer overall health [48]. However, they found no difference in mental components such as anxiety, emotional role limitation, and no significant differences in physical activity and work ability. Two other studies used the same instrument for HRQoL (SF-36) with a similar age group (mean age 49) and found similar results [5, 9]. They found that the physical health component had the biggest difference between ME/CFS and healthy control groups [9] or their caregivers [5]. For mental health, the mental health summary component had the smallest difference between ME/CFS and healthy control groups [5] and other disease group [9]. Moreover, Kingdon et. al. found employment and income were associated with loss of functional status [9], which was similar to our findings. However, due to the small study numbers, our study did not include employment status in the adjusted linear regression model. In our findings, ME/CFS symptom severity is the main negatively affecting factor for both lower physical and psychological health indices in all groups. This helps validate the use of PQ-Sym12 instrument.

### Strength and Limitations

This study included data from a large population-based cohort, making the data representative of the population of British Columbia at the age group studied. Our study also had good response rates, with 44% (85/195) of self-reported CFS participants completing all three surveys.

There were several limitations to our study that may have impacted our findings. First, the question posed to participants regarding their chronic fatigue diagnosis in the HLQ at the 2nd participant time-point was conducted by BCGP and not the study team; therefore, the study team had no influence on how the question was asked. The diagnosis of ME/CFS was not confirmed by physicians at the time of the 3^rd^ participant time-point. Second, as discussed above, people representing the elderly population in our study population were few. Moreover, our sample size did not have anyone younger than 35 years, meaning that younger age groups were not represented in our study. However, the observations of this study in the province of BC, with acceptable response and completion rates, indicating that this study is feasible for adoption of a further nation-wide study on ME/CFS.

### Implications

Our study showed that the use of self-reported diagnosis of ME/CFS is not a reliable indication to estimate the prevalence of this disease and is not well suited for clinical or epidemiological studies, due to the potential for bias as a result of misclassification of disease status. Participants were asked if they were willing to contribute to future storage of their data, participate in further research related to this study, and if they would like to be contacted for other research studies. Virtually all participants (98.9%) agreed to future storage of their data, further research related to this study, as well as future contact for other research studies. These participants could be included as part of an expansion of the first population cohort of individuals with confirmed ME/CFS to a national level to form a national Canadian ME cohort. We are currently planning a national population-based cohort study, which will enable the validation our findings and examine regional differences across Canada.

## CONCLUSIONS

This is the first population-based study on ME/CFS in British Columbia, Canada, which supports the planning and conduct of similar population-based cohort studies on the epidemiology of ME/CFS at a national level. Our study provides preliminary and exploratory findings on the epidemiology of ME/CFS based on self-reported previously diagnosed CFS individuals and their HRQoL and risk factors. Results gathered from this study have shown that the use of self-reported ME/CFS diagnoses may not be reliable and clinical assessment is necessary for diagnosis. They also confirm significant limitations in physical functioning and quality of life in people with ME/CFS. Further well-established data registries including all age groups across Canada would be beneficial for people affected by ME/CFS in terms of diagnosis, more accurate epidemiology, and for inclusion in treatment trials.

## Data Availability

All data produced in the present study are available upon reasonable request to the authors

## Acknowledgments

The authors would like to thank the BC Generations Project, in particular Jaclyn Parks and Jessica Chu, for their collaboration. The data used in this research were made available by the BC Generations Project. As well, the authors would like to thank the Women’s Health Research Institute: Nicole Prestley, manager research and knowledge translation for Interdisciplinary Canadian Collaborative ME (ICanCME) grant application, and Sabina Dobrer, senior statistician for her statistical advising. The authors would also like to thank the ICanCME Network and the Canadian Institutes of Health Research for providing funds to conduct the study. Dr. Nacul also would like to acknowledge and thank the Pacific Public Health Foundation (previously known as the BCCDC Foundation for Public Health) and BC Women’s Health Foundation for their sponsorship of his protected research time.

## REFERENCES

1. Nacul LC, Lacerda EM, Pheby D, Campion P, Molokhia M, Fayyaz S, et al. Prevalence of myalgic encephalomyelitis/chronic fatigue syndrome (ME/CFS) in three regions of England: a repeated cross-sectional study in primary care. BMC Medicine. 2011;9(1):91.

2. Lim E-J, Ahn Y-C, Jang E-S, Lee S-W, Lee S-H, Son C-G. Systematic review and meta-analysis of the prevalence of chronic fatigue syndrome/myalgic encephalomyelitis (CFS/ME). Journal of translational medicine. 2020;18(1):1–15.

3. Canada S. Canadian Community Health Survey, 2014 2014 [cited 2023 February 13]. Available from: https://www150.statcan.gc.ca/n1/daily-quotidien/150617/dq150617b-eng.htm https://www.ctvnews.ca/health/chronic-fatigue-syndrome-in-canada-even-worse-than-we-thought-survey-1.3539595?autoPlay=true.

4. Canada S. Canadian Community Health Survey, 2017 2017 [Available from: https://hdl.handle.net/11272.1/AB2/VXU9UQ/DL3LBC.

5. Nacul LC, Lacerda EM, Campion P, Pheby D, Drachler MdL, Leite JC, et al. The functional status and well being of people with myalgic encephalomyelitis/chronic fatigue syndrome and their carers. BMC Public Health. 2011;11(1):402.

6. Hardt J, Buchwald D, Wilks D, Sharpe M, Nix WA, Egle UT. Health-related quality of life in patients with chronic fatigue syndrome: an international study. J Psychosom Res. 2001;51(2):431–4.

7. Rakib A, White PD, Pinching AJ, Hedge B, Newbery N, Fakhoury WK, et al. Subjective quality of life in patients with chronic fatigue syndrome. Qual Life Res. 2005;14(1):11–9.

8. Falk Hvidberg M, Brinth LS, Olesen AV, Petersen KD, Ehlers L. The Health-Related Quality of Life for Patients with Myalgic Encephalomyelitis / Chronic Fatigue Syndrome (ME/CFS). PLoS One. 2015;10(7):e0132421.

9. Kingdon CC, Bowman EW, Curran H, Nacul L, Lacerda EM. Functional status and well-being in people with myalgic encephalomyelitis/chronic fatigue syndrome compared with people with multiple sclerosis and healthy controls. PharmacoEconomics-open. 2018;2(4):381–92.

10. Rasa S, Nora-Krukle Z, Henning N, Eliassen E, Shikova E, Harrer T, et al. Chronic viral infections in myalgic encephalomyelitis/chronic fatigue syndrome (ME/CFS). J Transl Med. 2018;16(1):268.

11. Lacerda EM, Geraghty K, Kingdon CC, Palla L, Nacul L. A logistic regression analysis of risk factors in ME/CFS pathogenesis. BMC Neurology. 2019;19(1):275.

12. Jain V, Arunkumar A, Kingdon C, Lacerda E, Nacul L. Prevalence of and risk factors for severe cognitive and sleep symptoms in ME/CFS and MS. BMC Neurol. 2017;17(1):117.

13. Cliff JM, King EC, Lee JS, Sepúlveda N, Wolf AS, Kingdon C, et al. Cellular Immune Function in Myalgic Encephalomyelitis/Chronic Fatigue Syndrome (ME/CFS). Front Immunol. 2019;10:796.

14. Rivas JL, Palencia T, Fernández G, García M. Association of T and NK Cell Phenotype With the Diagnosis of Myalgic Encephalomyelitis/Chronic Fatigue Syndrome (ME/CFS). Front Immunol. 2018;9:1028.

15. Bansal R, Gubbi S, Koch CA. COVID-19 and chronic fatigue syndrome: An endocrine perspective. J Clin Transl Endocrinol. 2022;27:100284.

16. Grabowska AD, Lacerda EM, Nacul L, Sepúlveda N. Review of the Quality Control Checks Performed by Current Genome-Wide and Targeted-Genome Association Studies on Myalgic Encephalomyelitis/Chronic Fatigue Syndrome. Frontiers in Pediatrics. 2020;8.

17. Conroy K, Bhatia S, Islam M, Jason LA. Homebound versus Bedridden Status among Those with Myalgic Encephalomyelitis/Chronic Fatigue Syndrome. Healthcare. 2021;9(2):106.

18. Mirin AA, Dimmock ME, Jason LA. Updated ME/CFS prevalence estimates reflecting post-COVID increases and associated economic costs and funding implications. Fatigue: Biomedicine, Health & Behavior. 2022;10(2):83–93.

19. Toogood PL, Clauw DJ, Phadke S, Hoffman D. Myalgic encephalomyelitis/chronic fatigue syndrome (ME/CFS): Where will the drugs come from? Pharmacological Research. 2021;165:105465.

20. Nacul L, Authier FJ, Scheibenbogen C, Lorusso L, Helland IB, Martin JA, et al. European Network on Myalgic Encephalomyelitis/Chronic Fatigue Syndrome (EUROMENE): Expert Consensus on the Diagnosis, Service Provision, and Care of People with ME/CFS in Europe. Medicina (Kaunas). 2021;57(5).

21. Bateman L, Bested AC, Bonilla HF, Chheda BV, Chu L, Curtin JM, et al. Myalgic Encephalomyelitis/Chronic Fatigue Syndrome: Essentials of Diagnosis and Management. Mayo Clinic Proceedings. 2021;96(11):2861–78.

22. Steiner S, Fehrer A, Hoheisel F, Schoening S, Aschenbrenner A, Babel N, et al. Understanding, diagnosing, and treating Myalgic encephalomyelitis/chronic fatigue syndrome – State of the art: Report of the 2nd international meeting at the Charité Fatigue Center. Autoimmunity Reviews. 2023;22(11):103452.

23. Dhalla A, McDonald TE, Gallagher RP, Spinelli JJ, Brooks-Wilson AR, Lee TK, et al. Cohort Profile: The British Columbia Generations Project (BCGP). Int J Epidemiol. 2019;48(2):377–8k.

24. Dummer TJB, Awadalla P, Boileau C, Craig C, Fortier I, Goel V, et al. The Canadian Partnership for Tomorrow Project: a pan-Canadian platform for research on chronic disease prevention. Cmaj. 2018;190(23):E710–e7.

25. Lacerda EM, Bowman EW, Cliff JM, Kingdon CC, King EC, Lee JS, et al. The UK ME/CFS Biobank for biomedical research on Myalgic Encephalomyelitis/Chronic Fatigue Syndrome (ME/CFS) and Multiple Sclerosis. Open J Bioresour. 2017;4.

26. Ware J, Kosinski M, Keller S. SF-36 physical and mental health summary scales. A user’s manual. 2001;1994.

27. Carruthers BM, Jain AK, De Meirleir KL, Peterson DL, Klimas NG, Lerner AM, et al. Myalgic Encephalomyelitis/Chronic Fatigue Syndrome. Journal of Chronic Fatigue Syndrome. 2003;11(1):7–115.

28. Committee on the Diagnostic Criteria for Myalgic Encephalomyelitis/Chronic Fatigue S, Board on the Health of Select P, Institute of M. The National Academies Collection: Reports funded by National Institutes of Health. Beyond Myalgic Encephalomyelitis/Chronic Fatigue Syndrome: Redefining an Illness. Washington (DC): National Academies Press (US) Copyright 2015 by the National Academy of Sciences. All rights reserved.; 2015.

29. StataCorp. Stata Statistical Software: Release 17. In: Station C, editor. TX: StataCorp LLC.; 2021.

30. Ware JE, Jr., Sherbourne CD. The MOS 36-item short-form health survey (SF-36). I. Conceptual framework and item selection. Med Care. 1992;30(6):473–83.

31. Hays RD, Sherbourne CD, Mazel RM. The RAND 36-Item Health Survey 1.0. Health Economics. 1993;2:217–27.

32. Lim E-J, Son C-G. Review of case definitions for myalgic encephalomyelitis/chronic fatigue syndrome (ME/CFS). Journal of Translational Medicine. 2020;18(1):289.

33. Brurberg KG, Fønhus MS, Larun L, Flottorp S, Malterud K. Case definitions for chronic fatigue syndrome/myalgic encephalomyelitis (CFS/ME): a systematic review. BMJ Open. 2014;4(2):e003973.

34. Christley Y, Duffy T, Martin CR. A review of the definitional criteria for chronic fatigue syndrome. Journal of Evaluation in Clinical Practice. 2012;18(1):25–31.

35. Jason LA, Brown A, Evans M, Sunnquist M, Newton JL. Contrasting chronic fatigue syndrome versus myalgic encephalomyelitis/chronic fatigue syndrome. Fatigue: biomedicine, health & behavior. 2013;1(3):168–83.

36. Bowen J, Pheby D, Charlett A, McNulty C. Chronic Fatigue Syndrome: a survey of GPs’ attitudes and knowledge. Family Practice. 2005;22(4):389–93.

37. Cairns R, Hotopf M. A systematic review describing the prognosis of chronic fatigue syndrome. Occupational medicine. 2005;55(1):20–31.

38. Nacul L, O’Boyle S, Palla L, Nacul FE, Mudie K, Kingdon CC, et al. How Myalgic Encephalomyelitis/Chronic Fatigue Syndrome (ME/CFS) Progresses: The Natural History of ME/CFS. Frontiers in Neurology. 2020;11.

39. Vercoulen J, Swanink C, Fennis J, Galama J, Van der Meer J, Bleijenberg G. Prognosis in chronic fatigue syndrome: a prospective study on the natural course. Journal of Neurology, Neurosurgery & Psychiatry. 1996;60(5):489–94.

40. Flo E, Chalder T. Prevalence and predictors of recovery from chronic fatigue syndrome in a routine clinical practice. Behaviour Research and Therapy. 2014;63:1–8.

41. Valdez AR, Hancock EE, Adebayo S, Kiernicki DJ, Proskauer D, Attewell JR, et al. Estimating Prevalence, Demographics, and Costs of ME/CFS Using Large Scale Medical Claims Data and Machine Learning. Front Pediatr. 2018;6:412.

42. Table 13-10-0849-01. Chronic conditions among seniors aged 65 and older, Canadian Health Survey on Seniors, two-year period estimates. [Internet]. 2022 [cited March 2023]. Available from: https://www150.statcan.gc.ca/t1/tbl1/en/tv.action?pid=1310084901.

43. Bayliss K, Riste L, Fisher L, Wearden A, Peters S, Lovell K, et al. Diagnosis and management of chronic fatigue syndrome/myalgic encephalitis in black and minority ethnic people: a qualitative study. Primary health care research & development. 2014;15(2):143–55.

44. Anderson JS, Ferrans CE. The quality of life of persons with chronic fatigue syndrome. The Journal of nervous and mental disease. 1997;185(6):359–67.

45. Schweitzer R, Kelly B, Foran A, Terry D, Whiting J. Quality of life in chronic fatigue syndrome. Social Science & Medicine. 1995;41(10):1367–72.

46. Solomon L, Nisenbaum R, Reyes M, Papanicolaou DA, Reeves WC. Functional status of persons with chronic fatigue syndrome in the Wichita, Kansas, population. Health and quality of life outcomes. 2003;1(1):1–10.

47. Brown MM, Bell DS, Jason LA, Christos C, Bell DE. Understanding long-term outcomes of chronic fatigue syndrome. Journal of clinical psychology. 2012;68(9):1028–35.

48. Bernhoff G, Rasmussen-Barr E, Bunketorp Käll L. A comparison of health-related factors between patients diagnosed with ME/CFS and patients with a related symptom picture but no ME/CFS diagnosis: a cross-sectional exploratory study. Journal of Translational Medicine. 2022;20(1):577.

