## Supplemental Figure 1 for "Epidemiology of Myalgic Encephalomyelitis among individuals with self-reported Chronic Fatigue Syndrome in British Columbia, Canada, and their health-related quality of life"

**SUPPLEMENTARY FIGURE**

***S1 Fig. Flowchart describing recruitment process*.**

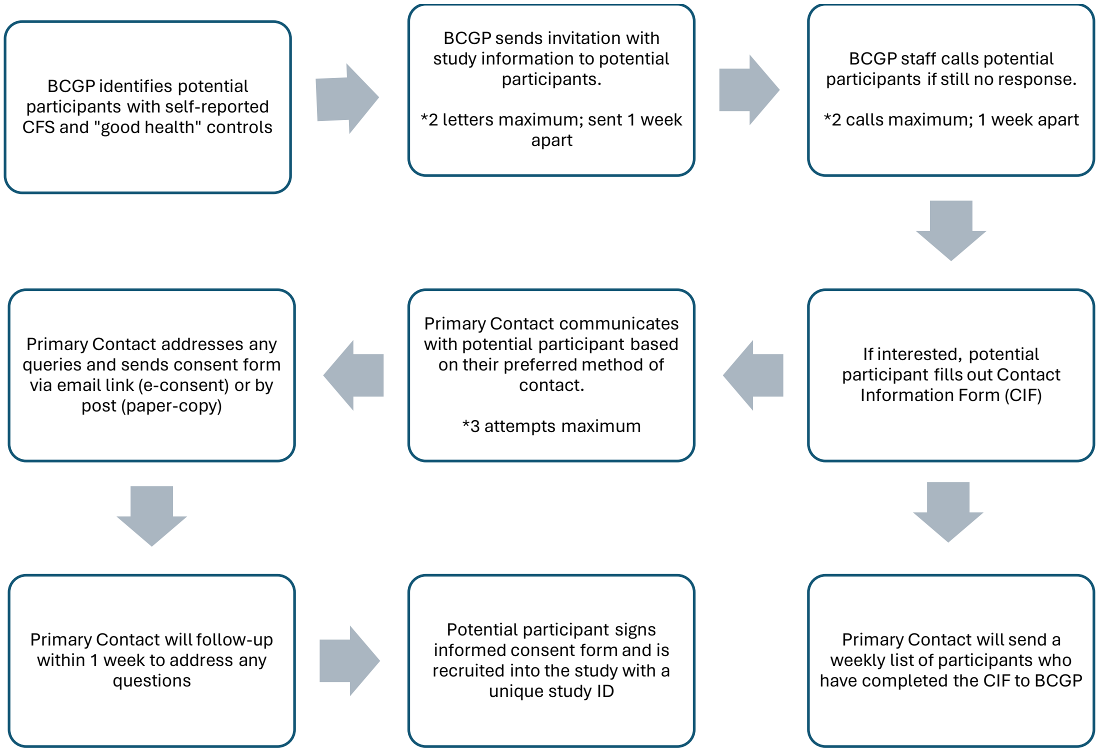


***Note:*** BCGP means British Columbia Generation Project cohort.

Primary Contact means a research assistant from our Research Team, who primarily contacted to all the participants in the study
